# Modeling Familial MASH by iPSC-Hepatocytes

**DOI:** 10.1101/2025.11.18.25338158

**Authors:** Cristina Esteva-Font, Jacquelyn J. Maher, Eric Rulifson, Mark Pabst, Nathan Bass, Holger Willenbring, Aras N. Mattis

**Author notes:** Contact Information: Aras N. Mattis, University of California San Francisco, 513 Parnassus Ave, HSW516, San Francisco, California 94143, USA.

## Abstract

Increasing obesity has led to a vast rise in metabolic dysfunction-associated steatotic liver disease (MASLD) in the general population with a significant fraction progressing to metabolic dysfunction-associated steatohepatitis (MASH). Patients with MASH progressively develop inflammation and fibrosis that over time can develop into cirrhosis with an increased risk for hepatocellular carcinoma. Despite the study of human hepatoma lines and development of numerous mouse models to dissect this disease, none of these truly function as a preclinical platform for the human disease. To faithfully model this disease pathogenesis, we identified several families with a genetic predisposition for MASH identified via the clinic, reprogramed their skin fibroblasts into induced pluripotent stem cells (iPSCs) and differentiated these to hepatocytes (iHeps). Focusing on one family, and compared to control iHeps, this MASH family showed increased baseline steatosis. Whole exome sequencing revealed the presence of numerous single nucleotide polymorphisms (SNP) of unclear significance. Interestingly these patients were heterozygous for the transmembrane 6 superfamily member 2 (TM6SF2) E167K SNP. We further analyzed these iHeps for spontaneous steatosis, apoptosis, mitochondrial function, and ER stress. Our findings illustrate the complexity of human genetic MASH patients but also highlights the power of using iHeps to characterize complex human diseases.

## Introduction

The increasing obesity of the modern era has led to rising metabolic dysfunction-associated steatotic liver disease (MASLD) prevalence with a significant fraction of these developing metabolic dysfunction-associated steatohepatitis (MASH) ^1^ that progressively leads to inflammation, fibrosis and cirrhosis with a significantly increased risk for hepatocellular carcinoma (HCC). We fully acknowledge that MASLD and MASH are indeed multifactorial and strongly impacted by environment, diet, and human genetics ^2,3^. From a patient perspective, starting with the discovery of patatin-like phospholipase domain-containing protein 3 (PNPLA3) SNP I148M^4^, many additional single nucleotide polymorphisms (SNPs) imparting risk for MASLD have been uncovered in genes including GCKR, TRIB1, G6P, and PPP1R3B^5^.

Many of these SNPs including PNPLA3 I148M have had complicated genetic phenotypes. TM6SF2 E167K was identified as strongly associated with steatosis, MASH, and even present in lean MASH. On the one hand, a wild-type E167 is a risk factor for increased serum triglycerides (TG) while the E167K SNP is protective against serum TGs but a risk factor for increased intrahepatic TGs^6^. As such, TM6SF2 E167K does affect an estimated 6.7%/4/7 of Caucasians/ Hispanics/ African Americans and represents a strong risk factor for MASLD. From a phenotype perspective, TM6SF2 E167K represents a clear phenotype risk factor and therefore a clear choice for characterization. To date only synthetic iPSC constructs containing TM6SF2 E167K and their derived induced hepatocytes have been reported^7^.

Despite the development of numerous mouse models to study and dissect this disease, given the large differences between mouse and human metabolism^8^, it is unclear if these model systems can be used to develop therapeutic treatments for human MASH. To circumvent genetic differences, and to develop a clearly translatable model system, we embraced familial MASH as a concept with strong genetics to dissect a multifactorial disease. This goes with that idea that there has been a general resurgence of studying families to understand complex diseases.

Modeling of human monogenetic diseases using induced pluripotent stem cells (iPSCs), has had significant successes investigating complex diseases including mitochondrial depletion syndrome, familial hypercholesterolemia, and APOB secretion phenotypes ^9–11^. We therefore embraced this methodology and took both a humble yet bold approach to model aspects of MASLD that were amenable to hepatocyte modeling.

To directly create relevant biologic models of MASH, we chose to use patients with the most severe form of the disease that have undergone liver transplant, as tissue donors to recreate this disease using induced pluripotent stem cell (iPSC) technology with differentiation into induced hepatocytes (iHeps). Because MASH is a complex genetic disease with environmental influence, we choose to focus on a single family with a genetic predisposition for MASH. By focusing on patients with this genetic tilt for MASH, we were able to establish iPSCs that when differentiated into iHeps, show both baseline differences in lipid accumulation as well as significant phenotypic accumulation of lipids and cell death during palmitate challenge. We went on to further characterize one of these lines looking at possible pathophysiologic mechanisms for human MASH in what is likely to involve multiple pathways.

## Materials and Methods

### Establishing patient fibroblast cell lines

Full-thickness, punch skin biopsies were taken according to published technique of from the volar aspect of the left non-dominant forearm^11^. Dermal fibroblasts were grown and frozen down at early passages for reprograming to iPSCs.

### Generation of iPSCs

iPSCs were generated using integration-free plasmids as previously described ^12^.

### Cell Culture of iPSCs

The media conditions to maintain human iPSCs have all been previously described ^13^.

### Whole Exome Sequencing

Whole exome sequencing was performed by Ambry Genetics using whole exome target enrichment, library preparation, 100 bp paired end processing on Illumina Hi Seq2000. A minimum of 100x raw coverage per exome was targeted with Human hg38 reference-guided alignment and analysis using the Ambry Variant Analyzer-clinical analysis tool.

### Induction of Definitive Endoderm (days 0-7)

Cells induced by recombinant human Activin A (100 ng/mL) Peprotech catalog #120-14E for 7 days at 20% oxygen, 5% C02. On the day before differentiation (day 0), cells were re-plated on Matrigel coated plates in the presence of 2µM doxycycline and 10µM Y-27632. On day 1 of induction, 2% KSR Gibco Catalog #10828028, CHIR-99021 Selleckchem catalog # S2924 (3mM), PI-103 (50nM) Selleckchem catalog # S1038 and recombinant human BMP4 (10ng/mL) Peprotech catalog #120-05ET and FGF2 (20ng/mL) Peprotech catalog #100-18B were added to the media. On day 2, 1% KSR, PI-103 (50nM) and BMP4 (10ng/mL) and FGF2 (20ng/mL) and on day 3, 0.2% KSR and PI-103 (50nM) were added to the media. Doxycycline hyclate (Acros Organics catalog #446061000) was added throughout definitive endoderm induction at various doses as described in text with a standard dose being 2μM.

### RNA isolation, Real-Time PCR analysis, and RNA Sequencing

Total RNA was isolated by Qiagen RNeasy mini kit or Zymo Direct-zol RNA kits. Complementary DNA (cDNA) was synthesized by qScript cDNA SuperMix (Quantabio). Quantitative Real-Time PCR (qrtPCR) was performed using FastStart Universal SYBR Green, Roche Diagnostics (Indianapolis, IN). RNA Sequencing was performed by BGI, Boston, MA. Poly-A RNA was selected using oligo-dT magnetic beads followed by N6 random priming.

### Statistical Analysis

Experiments were run in at least triplicate for each condition. Statistical significance was represented as standard error of the mean (SEM) and was determined using Student t test or Anova analysis as appropriate (GraphPad Prism, La Jolla, CA).

### Study Approval

All studies using human stem cell materials were carried out under approval by the University of California, San Francisco, Institutional Review Board (IRB) study approval 10-04393.

## Results

### Patients with a familial predisposition for MASH

As MASH is a complex disease with non-Mendelian genetics and is influenced by environmental factors, we chose to start with patients with a strong familial predisposition for the disease to increase the probability that we could potentially uncover phenotypes in vitro. The adult gastroenterology division at UCSF identified three families with increased predisposition among either 1^st^ degree relatives or among multiple generations (**Figure. 1B; Supplemental Fig. 1A**). We procured skin biopsies from patients from within each family. At least one patient in each family had a diagnosis of MASH and had undergone liver transplant at UCSF. Because patients without symptoms for MASH are not formally evaluated by the gold standard of liver biopsy, we could not safely use unaffected family members as potential controls.

**Figure 1.**
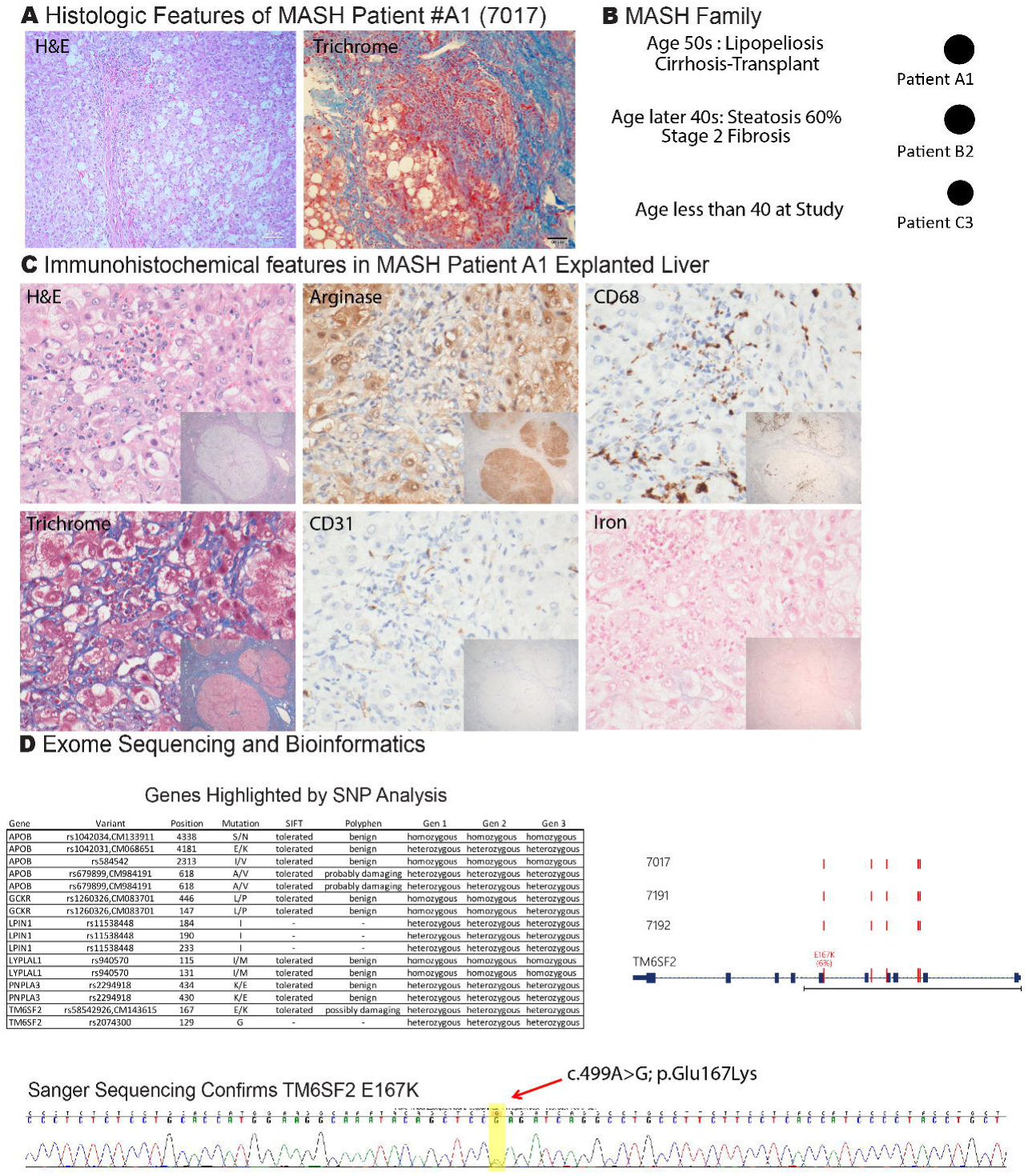
Evaluation of Familial MASH Family. (A) Histologic features of MASH patient A1 (cell line 7017) including H&E and trichrome stains showing severe steatosis, lipopeliosis, and cirrhosis (on Gomori trichrome). (B) Schematic of one MASH Family with patient A1 (cell line 7017) undergoing transplant in age 50s for cirrhosis due to MASH. Relative patient B2 (cell line 7191), and the third relative C3 (cell line 7192) were also clinically diagnosed with MASH. (C) Immunohistochemical and tissue-stained features in MASH Patient explanted liver including H&E, Arginase stain, CD68, Trichrome, CD31, and Iron stains. (D) Whole exome sequencing and Bioinformatics shows MASH genes highlighted by SNP analysis including presence of TM6SF2 E167K in all three generations confirmed by sanger sequencing.

Because one family had a strong history of early MASH and multiple generations that were available for skin biopsy as well as liver biopsy, we chose to focus our efforts on this family. The first subject with pseudonym A1, was a female of northern European ancestry who had undergone a liver transplant at UCSF in her 50s for subacute liver failure. The explanted liver revealed mixed micro-and macronodular cirrhosis with moderate inflammation and extensive steatosis with lipopeliosis, features compatible with cirrhosis secondary to MASH. To give a complete overview, we evaluated an extensive set of markers including Arginase (for hepatocytes, CD68 (macrophage), Trichrome (fibrosis), CD31 (vascular proliferation), Iron stain, PASD (activated macrophage), T4 collagen (fibrosis), and Factor 8 (functional staining) (**Figure 1A, 1C and 2A**). Other causes of liver disease, including viral hepatitis and autoimmune hepatitis were excluded through appropriate serological and histological testing, and there was no antecedent history of exposure to potentially hepatotoxic medications. She drank no alcohol either prior or subsequent to her transplant. Moderate iron accumulation was noted in the allograft during the first 2 years following the transplant associated with hyperferritinemia with a normal iron saturation index. Testing for HFE mutations C286Y, and H63D showed the presence of only wild-type alleles (data not shown), hence excluding classic genetic hemochromatosis. Iron staining of the explanted liver also showed rare non-parenchymal iron (**Figure 1C**). Other significant associated medical problems included obesity (5-year BMI range: 38.8 – 27.4), type 2 diabetes mellitus, hypertension, hypothyroidism, mild chronic kidney insufficiency associated with bilateral renal artery stenosis, mild cerebrovascular disease, dyslipidemia (triglyceride 692 mg/dl, Total cholesterol 200 mg/dl, LDLc 82 mg/dl, HDLc 36mg/dl), and hyperuricemia and gout. The second subject (B2), a relative of A1, was patient in their later 40s with morbid obesity (BMI=47) and co-morbidities including Type 2 diabetes mellitus and dyslipidemia. An earlier liver biopsy performed for elevated liver aminotransferase levels had shown a typical MASH histology. Other causes of liver disease had been excluded by appropriate serological testing and the patient did not drink alcohol. A more recent follow-up liver biopsy showed a classic pattern of advanced MASH with steatosis, and hepatocyte ballooning (both grade 2), inflammation and stage 2 fibrosis (**Figure 2B**). Skin fibroblast cell lines were established from both patients A1, B2 and a third younger relative in her 20s with suspected MASH (patient C3), with the first two having liver biopsy confirmed MASH.

**Figure 2.**
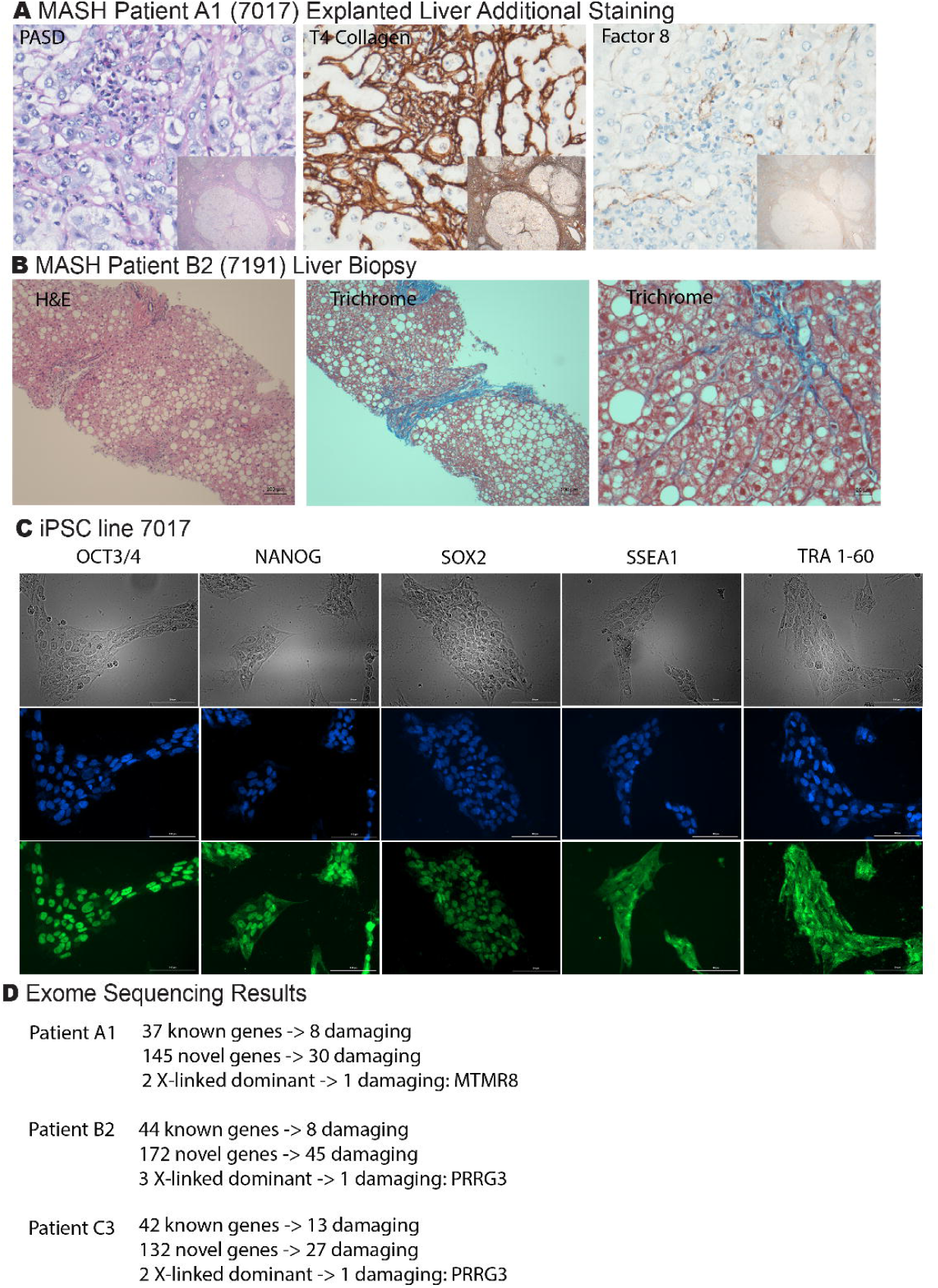
Evaluation of Familial MASH Family and Development of iPSCs. (A) Histologic features of MASH patient A1 (cell line 7017) including H&E, T4 Collagen, and Factor 8 immunohistochemical stains. (B) Histologic features of MASH patient B2 (cell line 7191) including H&E and trichrome stains showing severe steatosis, centrizonal pericellular fibrosis (Stage 2) on Gomori trichrome. (C) Brightfield and fluorescent stains showing markers of pluripotency including OCT3/4, NANOG, SOX2, SSEA1, and TRA 1-60 in iPSC line 7017. (Blue = Hoechst 33342; Green = GFP secondary antibody staining). (D) Whole exome sequencing results schematic including the number of novel and known genes.

While MASLD and MASH are considered complex diseases with complex genetics, ethnic predisposition towards MASH has been previously described ^4,6^. Ethnic predisposition has been further categorized to contain a preponderance of PNPLA3 in the case of Hispanic-Americans, and Caucasians while the Black population is relatively protected ^4^. We therefore confirmed the PNPLA3 status of our patient lines and found that none of them were positive for these genetic markers of predisposition (data not shown).

To further characterize these patients, we reprogrammed these patient fibroblasts to induced pluripotent stem cells (iPSCs) using both a lentiviral approach with the Yamanka factors including (Oct3/4, Sox2, Klf4, and L-Myc)^14^ as well as a plasmid based non-integrating DNA approach with an expanded set of genes including Oct3/4, p53 shRNA, Sox2, Klf4, L-Myc, and Lin28 ^12^. Delivery of the genes yielded iPSC and partially reprogramed colonies over 3-4 weeks that were subsequently subcloned into individual cell lines and confirmed for pluripotency (**Figure 2C, Supplemental Figure 1B**).

### Whole exome sequencing of MASH Family

In a bid to understand if there were novel mutations present within these patients and to better characterize their risk for MASLD/MASH, we performed whole exome sequencing (Ambry Genomics), see methods. As expected, a large number of mutations were identified by the Ambry Variant Analyzer (**Figure 2D**). The mutations included damaging homozygous and heterozygous genes listed in **supplemental Tables 1-5**. A curated set of interesting mutations is shown in **Figure 1D** including TM6SF2 E167K. Interestingly the patients also had a significant number of mutations in APOB. Given the importance of the TM6SF2 E167K mutation, we performed Sanger sequencing to confirm this mutation (**Figure 1D**).

### Quantitative phenotypes in MASH Patient iHeps

After establishing iPSCs from bonafide MASH patients, we set out to measure quantitative phenotypes that could confirm pre-existing hypotheses on MASH pathogenesis or uncover novel frameworks to think about the disease. First, we generated iPSC-hepatocytes (iHeps) using our previously published methodology^15^. Testing the iHeps in an unchallenged state, revealed increased baseline lipid accumulation in MASH patients (**Figure 3A** – Nile red staining for lipids). We next tested the response of familial MASH patients to palmitate measuring both cell viability and steatosis normalized per nucleus (**Figure 3B**). The findings show there was a decrease in MASH iHep cell viability compared to controls while there was a simultaneous increase in steatosis in response to palmitate challenge.

**Figure 3.**
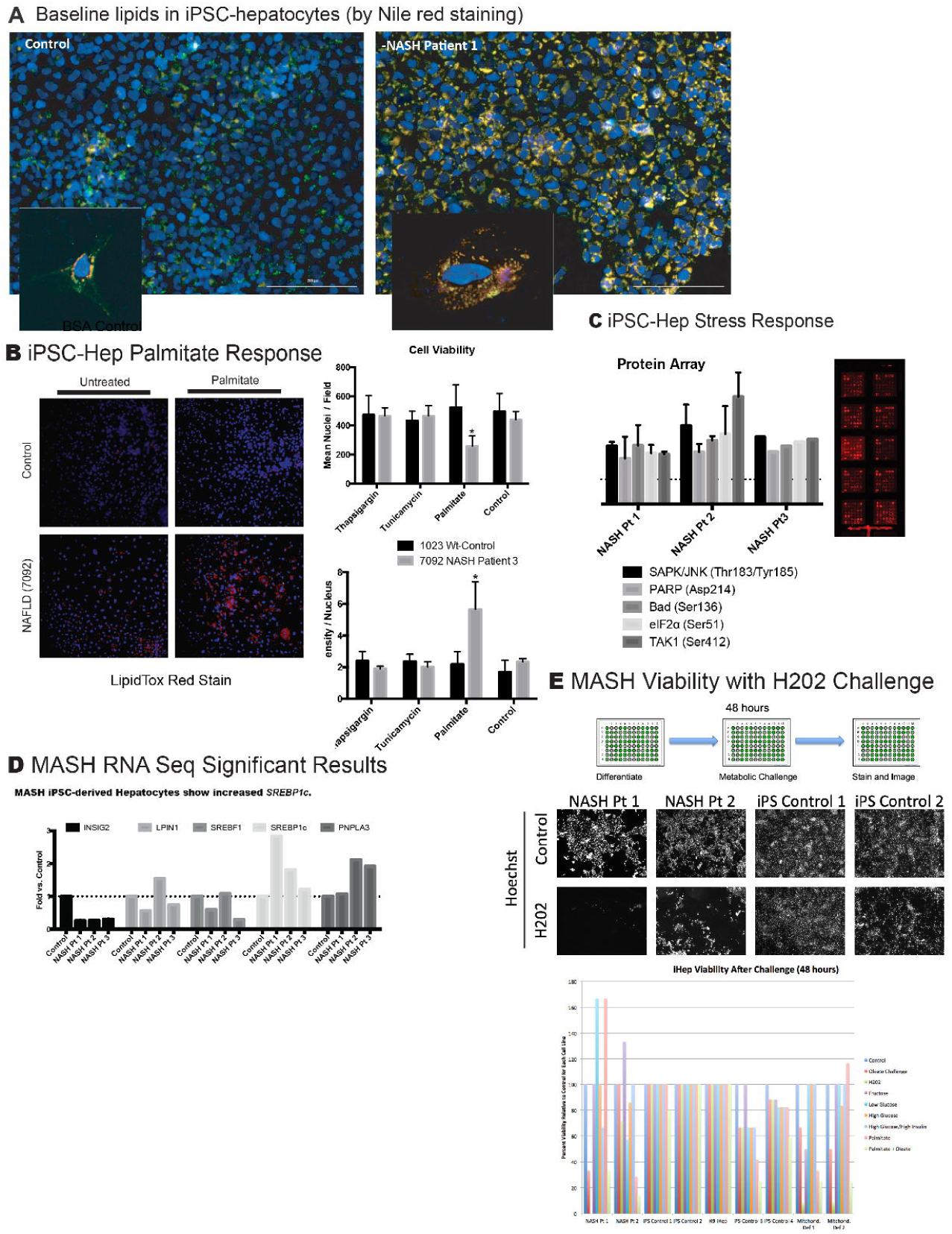
Familial MASH iHeps Reveal Underlying Metabolic Defects. (A) Baseline lipid accumulation in control iHeps (cell line 1023) versus MASH Patient A1 iHeps (cell line 7017) by fluorescent Nile Red staining (merged fluorescence from red and green channels) (Blue = Hoechst 33342 nuclear staining) scale bar = 100µM, (n=6). MASH Patient A1 shows significant baseline lipid accumulation. Inset shows single cell with lipid staining. (B) iPSC-Hepatocyte palmitate challenge in control (cell line 1023) versus MASH (cell line 7092) (500µM palmitate) for 24 hours with subsequent lipid staining (LipidTox Red; Blue = Hoechst 33342 nuclear staining) and quantification by Cytation5 imaging. Steatosis was also tested in palmitate versus Thapsigargin, Tunicamycin and control (DMSO). Mean nuclei per field were quantified for cell viability and lipid density per nucleus were quantified for steatosis (n=4). (C) iPSC-Hepatocyte Stress response signaling pathways array. MASH patients 001, 002, and 003 iHep cell extracts were processed with Cell Signaling Stress Response Pathway array. The results show significant activation of stress pathways versus control (dotted line – cell line 1023) (n=2). (D) MASH iHep RNA Sequencing Significant results show activation of de novo lipogenesis pathways with *SREBP1c* increased transcription with simultaneous repression of *INSIG2* pathway. (E) MASH iHep viability after H_2_0_2_ challenge shows that MASH patient A1 and B2 iHeps are much more sensitive than control iHeps to H_2_0_2_ challenge after 48 hours. We also performed oleate challenge, fructose challenge, glucose challenge and palmitate challenge.

Increased cell death suggested increased cellular ER stress and therefore we made protein extracts from MASH iHeps and tested phosphorylated protein activation via a protein array testing for numerous pathway activation including SAPK/JUNK, PARP, Bad, eIF2α and TAK1. The data shows that many ER stress pathways were activated at least 2-fold above baseline (**Figure 3C** – dotted line is normalized at 1).

To perform a pathway evaluation, we performed RNA sequencing and looked at a targeted analysis of pathways related to lipid metabolism comparing MASH iHeps from MASH patient A1 (7017), MASH patient B2 (7191), and MASH C3 (7192) to control lines including 1023 and others (**Supplemental Table 6**). In our MASH study, we found that SREBP1c was always upregulated with INSIG2 being downregulated suggesting that the cells were upregulating de-novo lipogenesis while downregulating the protein break on de-novo lipogenesis (**Figure 3D**). Interestingly in 2 of 3 MASH iHep patients, the levels of PNPLA3 were upregulated despite no amino acid changes at PNPLA3 148.

Finally, we challenged our cells using a hydrogen peroxide viability challenge over 48 hours. We noted that our MASH patient iHeps were significantly unable to sustain viability in face of the H202 challenge compared to most of our control lines suggesting impaired ability to deal with increased stress (**Figure 3E**).

## Discussion

MASLD and MASH as well as other metabolic liver diseases have been difficult to model in traditional cell culture models especially since many of the human hepatoma models contain unique mutations including PNPLA3 148M. Isolation and modeling of hepatocytes and organoids from Bonafide MASH patients with varying severity (over multiple stages of fibrosis up to cirrhosis and transplant) is important to characterize the range of cellular responses, something which is currently very difficult to measure in humans. To date only iPSC lines have been developed where TM6SF2 E167K was edited into a normal so called “wild-type line”.

It will be critical to fully characterize other cellular compartments within these MASH patient lines, and to perform careful measurements that might uncover mechanistic insights into novel therapeutic target pathways. For example, it is phenotypically interesting that patient 7017 (patient A1) characterized in our studies had elevated serum lipids. While carrying of the TM6SF2 E167K would predict little secretion of Triglycerides from the liver, unless the liver was already saturated with lipids and overflowing into the serum, make the findings even more interesting. We look forward to using these cell lines as a platform for interrogation and discovery.

## Data Availability

All data produced in the present work are contained in the manuscript.

## Conflicts of Interest

Authors have nothing to disclose.

## Financial Support

H. Willenbring was supported by CIRM grant RN2-00950 and NIH grant P30 DK26743. A. N. Mattis was supported by CIRM grant TG2-01153 and NIH grants K08 DK098270, R01DK130391, R01ES033617, RC2DK136052, and R01DK132129. J. J. Maher was supported by NIH grants R01 DK068450 and P30 DK026743.

---

**Supplemental Figure 1:**
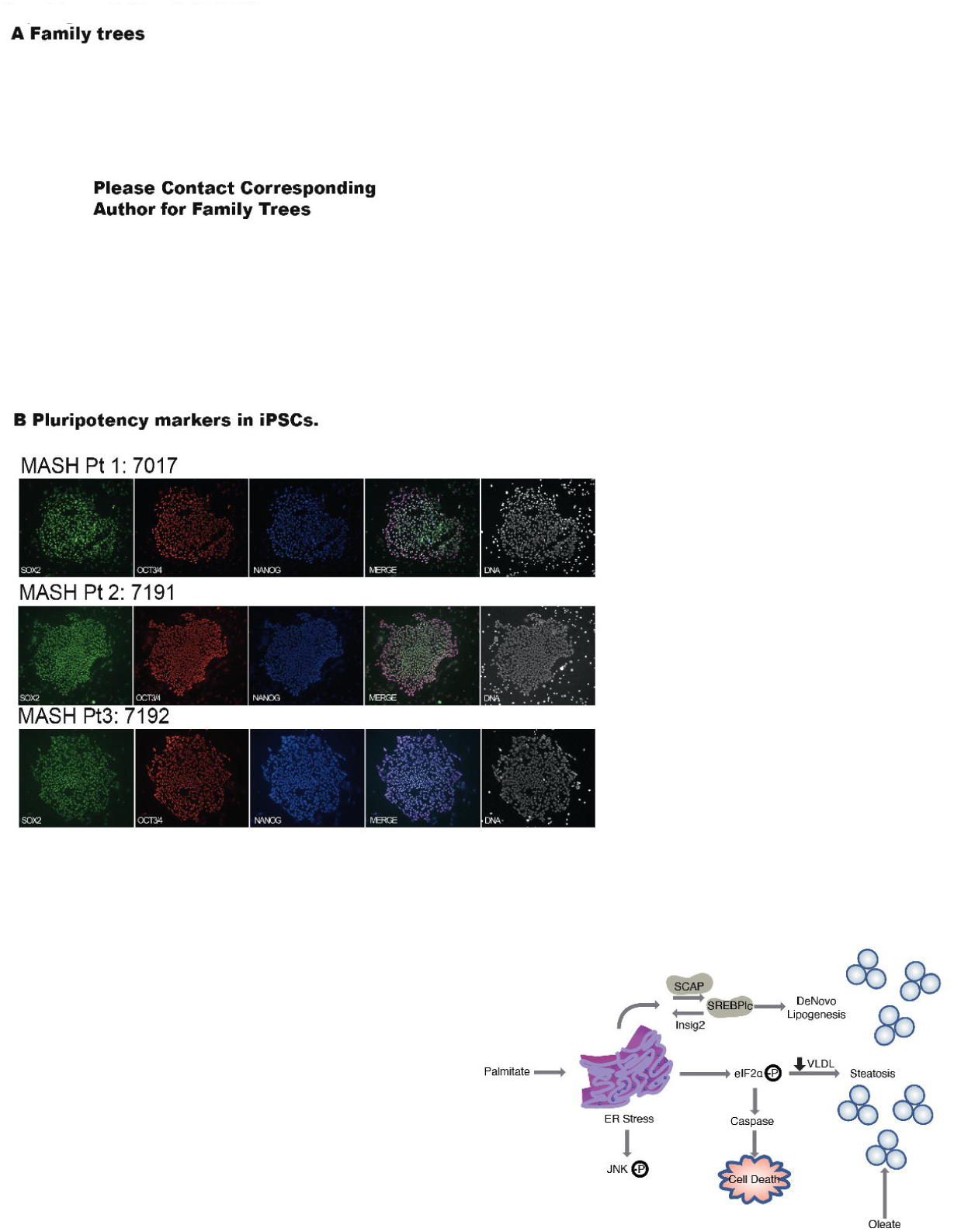
Familial MASH Families and iPSCs. (A) Three available family trees of Familial patients identified at our institution. Patients with a four-digit number underwent a skin biopsy that was used to develop iPSC lines. *Please contact corresponding author for family trees*. (B) Family B iPSC lines were confirmed by immunohistochemical staining for SOX2, OCT3/4, NANOG an merged photos including cell lines 7017, 7191, and 7192. (C) A model figure for de novo lipogenesis is shown.

**Supplemental Figure 2:**
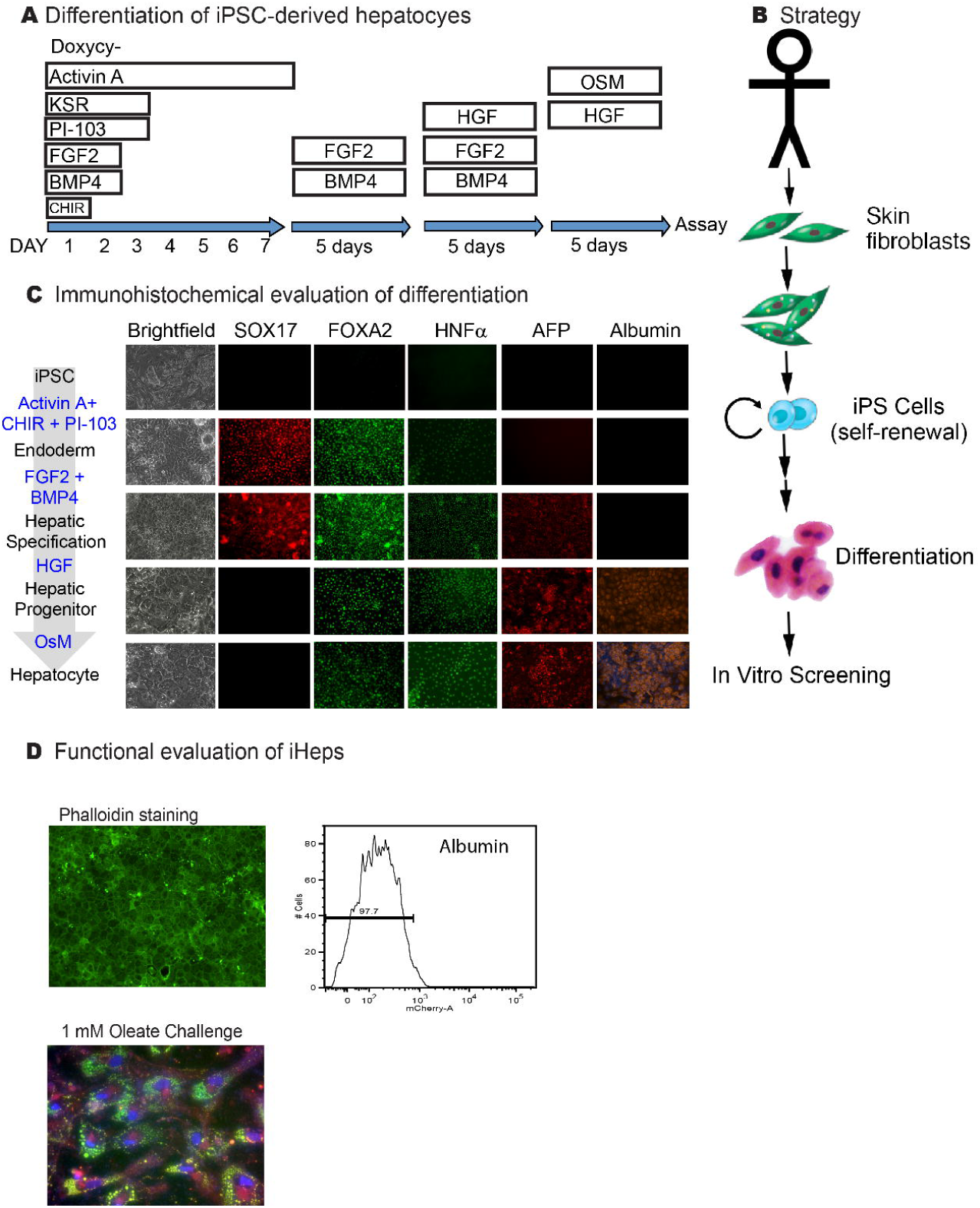
MASH iPSC Hepatocytes. (A) Differentiation of MASH iPSC-derived hepatocytes by our previously published methodology before assay over 23 days[Peaslee, 2021 #15]. (B) Overall strategy of taking patient skin fibroblasts to develop iPSCs for differentiation into iHeps that are then assayed by in vitro screening. (C) Immunohistochemical evaluation of differentiation for SOX17, FOXA2, HNF4α, AFP, and Albumin shows that iHeps mimic immature human hepatocytes after differentiation. (D) Functional evaluation of iHeps is shown including Phalloidin staining, Albumin staining by flow cytometry showing a high percentage of Albumin positive cells and finally a 1mM Oleate Challenge and Nile red staining.

## Notes

### Competing Interest Statement

Aras Mattis is a consultant for BioMarin, HepaTx, and Regeneron.

### Funding Statement

The study was funded by: H. Willenbring was supported by CIRM grant RN2-00950 and NIH grant P30 DK26743. A. N. Mattis was supported by CIRM grant TG2-01153 and NIH grants K08 DK098270, R01DK130391, R01ES033617, RC2DK136052, and R01DK132129. J. J. Maher was supported by NIH grants R01 DK068450 and P30 DK026743.

### Author Declarations

IRB of the University of California San Francisco gave ethical approval for this work.

